# Retrospective Evaluation of a Generative AI-Enabled Electronic Medical Record System in Primary Health Care Facilities in Kenya

**DOI:** 10.1101/2025.09.05.25335163

**Authors:** Ambrose Agweyu, Paul Mwaniki, Wilkister Musau, Robert Korom, Lynda Isaaka, Conrad Wanyama, Sarah Kiptinness, Najib Adan, Mira Emmanuel-Fabula, Bilal A. Mateen

**Affiliations:** KEMRI-Wellcome Trust Research Programme, Kenya; Keprecon, Kenya; London School of Hygiene and Tropical Medicine, United Kingdom; PATH, Kenya; Penda Health, Kenya; FATH, Switzerland; PATH, United Kingdom; University of Birmingham, UK

**Keywords:** Artificial Intelligence, Large Language Model, Primary Health Care, Kenya, Clinical Decision Support System

## Abstract

We conducted a retrospective evaluation of an electronic medical record-embedded large language model (LLM) clinical decision support system deployed across 16 primary care clinics in Kenya, between July-September 2024. A panel of trained physicians reviewed 1,469 records. Hallucinations were uncommon (50/1,469; 3.4%), most often involving mis-expanded acronyms or drug names. Clinical management guidance aligned with local guidelines in almost all cases (approximately 100%). Despite this, clinicians did not modify documentation in 62% of encounters. Safety assessments identified actively harmful recommendations from the LLM in 7.8% of encounters, with 67 such recommendations appearing in the final documentation. Conversely, risk present in the clinician’s initial notes was fully mitigated in 118 encounters (8.0% overall; 12.1% of amended cases). Overall, the tool showed strong potential to support quality improvement, but the asymmetric adoption of harmful versus beneficial outputs underscore the need for usability optimization, local guardrails, and prospective trials to confirm patient-level benefit.

## Introduction

An adequately trained and equitably distributed health workforce is essential for delivering high-quality care and averting preventable morbidity and mortality ^1^. However, many countries continue to face staffing shortages. The shortfall is most acute in sub-Saharan Africa, where reliance on cadres with short training pathways, high turnover, and sustained outmigration undermines service delivery ^2^. These workforce limitations give rise to wide disparities in quality of care across settings, and are compounded by frontline providers, particularly in marginalized areas, lacking access to structured clinical mentorship that might have otherwise mitigated the impact on care ^3^. Large language model (LLM)-based clinical decision support systems (CDSS) have the potential to help bridge these gaps by providing frontline clinicians with real-time, specialist physician-grade, contextually relevant diagnostic and management suggestions, potentially reducing clinically significant errors, avoiding unnecessary treatments, and minimizing delays in care by supporting appropriate and timely referrals ^4,5^.

Although evidence of the safety and contextual appropriateness of LLM-based CDSS in low-resource primary health care settings remains scarce ^6^, several studies in high-income countries have demonstrated the potential of this technology to improve diagnostic reasoning amongst primary care providers ^7,8^, support treatment planning ^9^, and even deliver therapeutic interventions ^10^. However, evidence from well-resourced settings is of limited value to decision makers in low- and middle-income countries (LMIC) because, as previous reports have noted ^11–14^, many of these models are trained predominantly on data that fail to account for local epidemiology, resource availability, and cultural context, reducing their reliability in LMIC health care settings. In the rare examples of proprietary models performing reasonably well on LMIC-based benchmarking datasets ^15^, the limited transparency into the underlying training data undermines the ability to draw meaningful insights into the true generalizability of the tool across settings, and there is no consensus on whether these in silico evaluations provide any actual ‘guarantees’ around real-world performance ^16^. In both cases, there is a very real risk wherein ‘naïve’ application of an LLM to a novel low-resource setting means that errors arising from the aforementioned biases in the training data might lead to flawed clinical reasoning or guidance and potentially compromise patient safety if taken at face value ^17^. Moreover, a recent user-experience research study in Kenya highlighted how this issue of lack of local grounding or outputs impacts frontline health worker perceptions of AI-based solutions, with clinicians reporting that poorly contextualized responses (e.g., suggesting a drug that was either unavailable or not part of the local protocol) undermined their confidence in the technology ^18^.

Recognizing these risks, the World Health Organization has emphasized that while AI could help “bridge gaps” in care for underserved communities, robust safeguards – such as local, real-world evaluations prior to deployment at scale – are needed to minimize risks and ensure ethical, context-appropriate use ^19^.

We sought to evaluate the quality and safety of LLM-generated clinical advice and its influence on clinician decision-making in the context of a network of 16 outpatient facilities operated by Penda Health, a private social enterprise serving urban communities in Nairobi and Kiambu counties, Kenya. Penda Health operates Level 3a and 3b facilities, which are 24/7 outpatient urgent and primary care centers staffed primarily by clinical officers (non-physician health care providers licensed to independently diagnose, prescribe, and manage a wide range of outpatient conditions). All of Penda Health’s facilities utilize a cloud-based electronic health record system (developed by Easy Clinic [India]), into which an LLM-based CDSS (Figure 1) was integrated and made available to staff.

**Figure 1:**
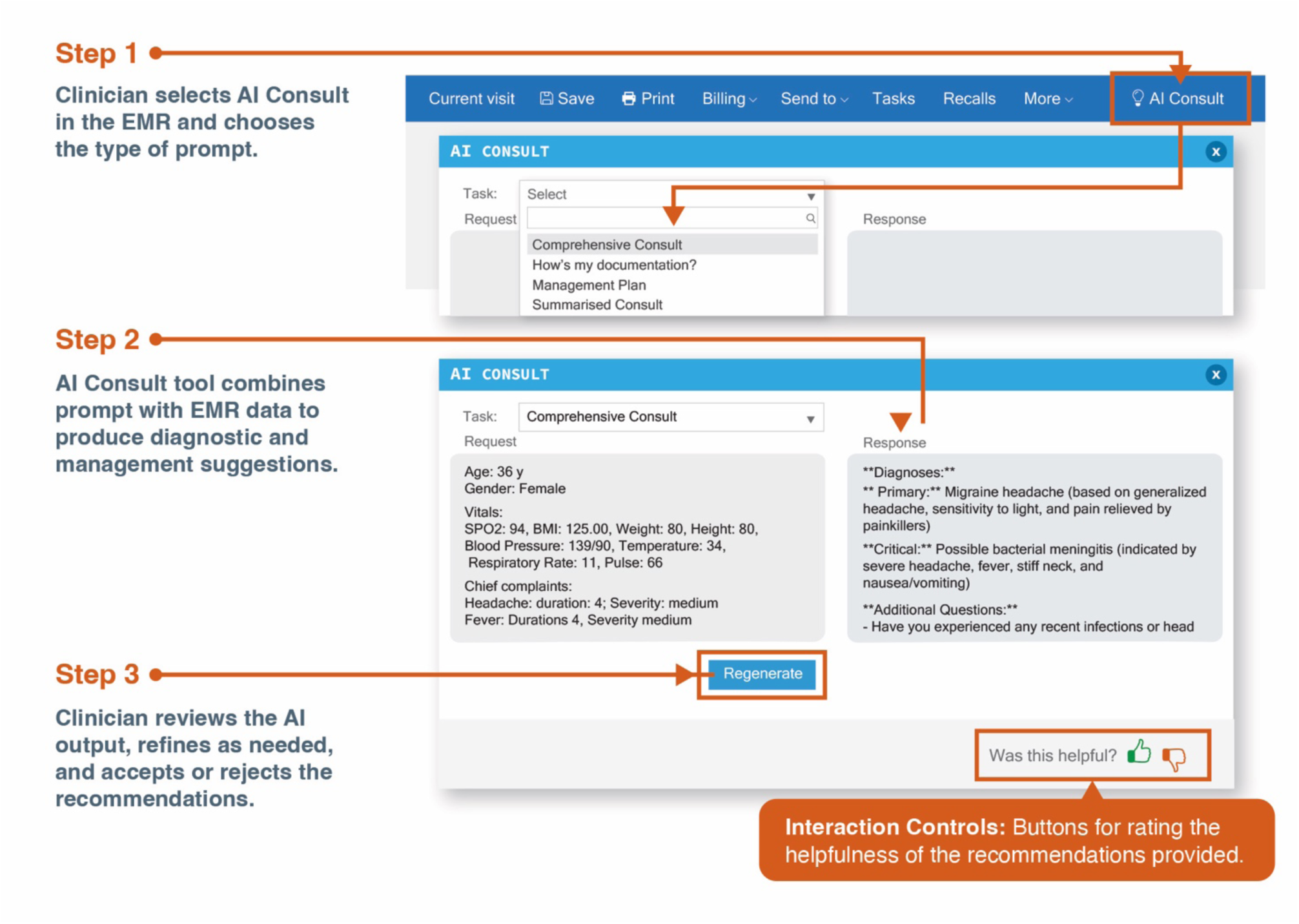
A Summary of the AI Consult V1 Workflow. Caption: The figure illustrates the GenAI-enabled workflow (referred to as AI Consult V1) that clinicians had access to during the study period. In summary, a button inside the electronic medical record (EMR) workflow allowed clinicians to select from one of four system-level prompts, which would be appended with all de-identified clinical data from the visit to create a query: · Comprehensive Consult: This prompt provided several paragraphs of feedback regarding differential diagnosis, diagnostic testing, history and physical examination gaps, and guideline-based treatments. Only encounters using this prompt were assessed in this study (see Supplementary Box 1 for the full system prompt). · Summarized Consult: The prompt provided no more than a paragraph of feedback, focusing on actionable steps a clinician could take to improve the quality of care. · Management Plan: The prompt provided a management plan for the condition without focusing on other aspects of care. · How’s my documentation?: This prompt provided a rating and constructive feedback points based on whether the clinical notes documented were adequate for the condition at hand. Notably, it was only introduced in September 2024. The prompt and EMR data (i.e., the query) would flow to the OpenAI Application Programming Interface (API), ChatGPT 4o, and return structured feedback to the clinician in a text box in real-time. Clinicians then had the option to either disregard the LLM output or consider it when making their clinical decisions. Clinical officers in the study retained full authority for their clinical decisions throughout the study. This figure and elements of the caption were originally published in Obong’o et al. ^18^, and reused under the associated CC-BY license provisions, without any amendments

## Results

### Study Cohort Characteristics

A total of 1,469 patient encounters were included in the evaluation, representing 1.9% of the 78,366 clinical consultations at the 16 Penda Health facilities over the three-month study period. Approximately a quarter of the patients were below 5 years of age (n = 350), 15% were aged 5-17 years (n = 225), half were aged 18-39 years (n = 730), 10% were aged 40-59 years (n = 150), and 1% were 60 years and above (n = 14). Based on the gender recorded in the EMR, 612 (42%) were male, and 857 (58%) were female. Key results stratified by patient age group are presented in Table 1.

**Table 1:** Selected Evaluation Metrics Across all Domains of Quality and Safety, Stratified by Patient Age.

| Characteristic | Overall<br>N = 1,469 | 0-4<br>N = 350 | 5-17<br>N = 225 | 18-39<br>N = 730 | 40-59<br>N = 150 | 60+<br>N = 14 |
| --- | --- | --- | --- | --- | --- | --- |
| Quality of initial clinical documentation |  |  |  |  |  |  |
| Totally inadequate/Needs significant improvement | 341 (23%) | 72 (21%) | 45 (20%) | 178 (24%) | 41 (27%) | 5 (36%) |
| Acceptable/High quality | 1,128 (77%) | 278 (79%) | 180 (80%) | 552 (76%) | 109 (73%) | 9 (64%) |
| Safety concerns associated with initial documentation |  |  |  |  |  |  |
| No safety concern identified | 922 (63%) | 212 (61%) | 130 (58%) | 490 (67%) | 82 (55%) | 8 (57%) |
| Mild/moderate concern identified | 410 (28%) | 114 (33%) | 82 (36%) | 164 (22%) | 46 (31%) | 4 (29%) |
| Severe or life-threatening risk identified | 137 (9.3%) | 24 (6.9%) | 13 (5.8%) | 76 (10%) | 22 (15%) | 2 (14%) |
| LLM addressed clinical issues identified in initial documentation | 1,419 (97%) | 343 (98%) | 215 (96%) | 700 (96%) | 147 (98%) | 14 (100%) |
| LLM prioritized the advice it provided appropriately based on initial documentation | 1,456 (99%) | 349 (100%) | 224 (100%) | 720 (99%) | 149 (99%) | 14 (100%) |
| LLM response had hallucinations | 50 (3.4%) | 10 (2.9%) | 14 (6.2%) | 21 (2.9%) | 5 (3.3%) | 0 (0%) |
| LLM effectively communicated | 1,459 (99%) | 349 (100%) | 224 (100%) | 723 (99%) | 149 (99%) | 14 (100%) |
| LLM response contained novel diagnostic insights |  |  |  |  |  |  |
| LLM provided novel contribution | 1,384 (94%) | 329 (94%) | 207 (92%) | 689 (94%) | 145 (97%) | 14 (100%) |
| LLM did not provide novel insight or gave misleading information | 85 (5.8%) | 21 (6.0%) | 18 (8.0%) | 41 (5.6%) | 5 (3.3%) | 0 (0%) |
| LLM response aligned with patient's context (epidemiology) | 1,462 (100%) | 349 (100%) | 223 (99%) | 727 (100%) | 149 (99%) | 14 (100%) |
| LLM provided clinical management advice | 1,460 (99%) | 347 (99%) | 224 (100%) | 727 (100%) | 149 (99%) | 13 (93%) |
| LLM response aligned with local clinical guidelines | 1,455 (99%) | 347 (99%) | 223 (99%) | 724 (99%) | 147 (98%) | 14 (100%) |
| LLM provided additional management-related insights that were not explicitly present in the initial documentation | 1,461 (99%) | 350 (100%) | 222 (99%) | 725 (99%) | 150 (100%) | 14 (100%) |
| Appropriateness of LLM response in patient's socioeconomic/cultural context or resource limitations |  |  |  |  |  |  |
| AI attempted to, unsuccessfully, adapt to patient's context | 2 (0.1%) | 0 (0%) | 1 (0.4%) | 1 (0.1%) | 0 (0%) | 0 (0%) |
| AI failed to acknowledge or did not attempt to adapt to patient's context | 2 (0.1%) | 1 (0.3%) | 0 (0%) | 1 (0.1%) | 0 (0%) | 0 (0%) |
| AI provided a response that was not culturally/contextually appropriate | 3 (0.2%) | 2 (0.6%) | 1 (0.4%) | 0 (0%) | 0 (0%) | 0 (0%) |
| No limitations or constraints described that required the LLM to adapt its response | 1,153 (78%) | 281 (80%) | 168 (75%) | 579 (79%) | 116 (77%) | 9 (64%) |
| Yes, AI adapted response fully to context | 309 (21%) | 66 (19%) | 55 (24%) | 149 (20%) | 34 (23%) | 5 (36%) |
| LLM response contains actively harmful recommendations | 115 (7.8%) | 24 (6.9%) | 20 (8.9%) | 56 (7.7%) | 15 (10%) | 0 (0%) |
| Initial Documentation Risk Mitigated by LLM |  |  |  |  |  |  |
| No | 548 (37%) | 141 (40%) | 96 (43%) | 240 (33%) | 65 (43%) | 6 (43%) |
| No risk was identified in the initial documentation | 803 (55%) | 182 (52%) | 110 (49%) | 431 (59%) | 73 (49%) | 7 (50%) |
| Yes | 118 (8.0%) | 27 (7.7%) | 19 (8.4%) | 59 (8.1%) | 12 (8.0%) | 1 (7.1%) |
| Harmful LLM output reflected in final documentation<sup>1</sup> | 67 (4.6%) | 16 (4.6%) | 13 (5.8%) | 25 (3.4%) | 13 (8.7%) | 0 (0%) |
| LLM response was helpful | 1,256 (86%) | 307 (88%) | 189 (84%) | 617 (85%) | 131 (87%) | 12 (86%) |

<sup>1</sup> Applicable in observations where LLM provided actively harmful response.

Respiratory system presentations were the most common (n = 561; 38%), followed by gastrointestinal (n = 362; 25%), and then genitourinary/reproductive (n = 217; 15%) presentations. Other categories included dermatological (n = 149; 10%), musculoskeletal (n = 130; 9%), febrile/infectious (n = 109; 7%), neurological/psychiatric (n = 73; 5%), ear, nose and throat/dental/ophthalmological (n = 64; 4%), cardiovascular (n = 39; 3%), and unspecified/other (n = 101; 7%).

Use of the AI-enabled CDSS increased over the evaluation period, although with substantial variation across clinics. In July 2024, consultations using the tool ranged from 29.3% (Zimmerman) to 53.7% (Kawangware). By September 2024, usage had increased across nearly all clinics, with 7/16 exceeding 60% utilization. The highest proportions were observed in Kangemi (80.6%), and Kasarani (75.1%). Clinics such as Zimmerman and Umoja 2 showed a more modest increase (reaching 44.7% and 40.7%, respectively, by September) (Supplementary Figure 1).

### Baseline Assessment of the Clinicians’ Input (To the AI Consult) Documentation

Over three-quarters of the cases assessed had initial clinical documentation rated as acceptable (n = 878; 60%) or of high quality (n = 250; 17%), while 279 records (19%) required significant improvement, and 62 (4%) were rated as totally inadequate. Inter-rater reliability statistics for all metrics are reported in Supplementary Table 1.

Safety concerns were identified in the initial documentation of 547 (37%) records. The most common safety concerns were inappropriate medication (272; 49%), omission of critical differential diagnoses (230; 42%), and incorrect diagnoses (120; 21.9%). Less frequent issues included incorrect or unsafe dosing (61; 11.2%), inappropriate investigations or tests (46; 8.4%), culturally or contextually inappropriate guidance (5; 0.9%), and other concerns (66; 12.1%). The perceived likelihood of harm associated with these concerns varied considerably: low in 243 cases (44%), moderate in 211 cases (39%), guaranteed in 42 cases (7.7%), and none in 51 cases (9.3%). Similarly, the severity of potential harm was rated as moderate in 212 cases (39%), mild in 198 (36%), severe in 112 (20%), and life-threatening in 25 (4.6%). Safety concerns were less common when initial documentation was high quality and rose as quality declined. Specifically, 14% (36/250) of high-quality notes prompted safety concerns, compared with 35% (309/878) for acceptable, 58% (163/279) for needs-significant-improvement, and 63% (39/62) for totally inadequate documentation, suggesting a stepwise gradient. In lower-quality strata, concerns also shifted toward higher likelihood and severity categories. Summaries of the likelihood, severity, and type of harm, stratified by each documentation quality rating, are presented in Supplementary Tables 2-6 and illustrated in Figure 2.

**Figure 2:**
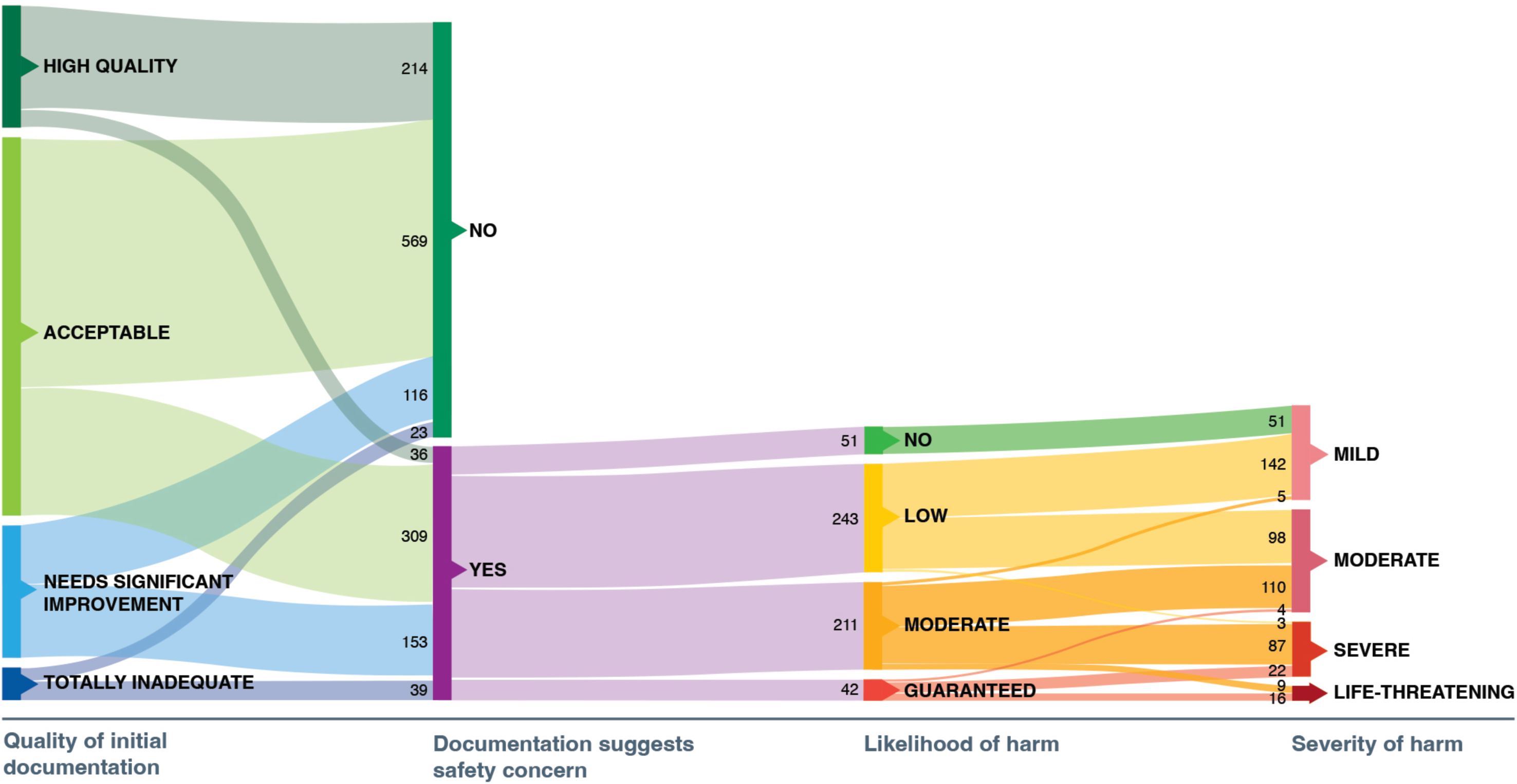
Sankey Plot of Documentation Quality, Associated Risk, Likelihood, Severity, and Type of Harm.

### Appropriateness of the LLM-based CDSS Outputs

#### Global Assessment

The majority of the AI-generated responses (1,213; 83%) fully addressed the clinical issues identified in the initial documentation, with a further 14% (n=206) doing so mostly. Only 3.3% (n=49) partially addressed the issues, and just one instance (<0.1%) showed no relevance to the clinical problem. Moreover, nearly all AI-generated responses (1,456; 99%) prioritized the advice appropriately based on the initial documentation. And finally, the majority of AI-generated responses (1,459; 99%) were assessed as having effectively communicated their advice.

Hallucinated content, defined as fabricated or clinically inaccurate information, was identified in 50 responses (3.4%). Post hoc qualitative review of evaluators’ comments grouped hallucinations into the following four themes: misidentified drugs or mis-expanded acronyms (n=14) (e.g., “FGC” read as “female genital circumcision” rather than fair general condition); contradictions of the initial documentation (n=8) (e.g., stating there were no signs of dehydration when the clinical note did not document hydration status); clinical parameter misinterpretation (n=8) (e.g., flagging SpO₂ 95% at Nairobi’s altitude ∼1,800m as abnormal); and, a category where the rationale for hallucination was insufficiently specified (n=20).

#### Diagnostic Reasoning

The presence of any diagnostic reasoning by the LLM was noted in nearly all outputs (1,443; 98%). Regarding the quality of differential diagnosis, 83% (1,220) of responses demonstrated strong and well-reasoned consideration of alternatives (or rightly affirmed that the clinician’s differential was appropriate), while 15% (223) had partial or incomplete differential reasoning (for example, demonstrated by a five-year-old child with a month-long history of rhinorrhea and snoring, focusing on sinusitis management without considering adenoidal hypertrophy or other obstructive etiologies). A small number of responses (24; 1.6%) contained major reasoning gaps or misleading information, and only two responses (0.1%) were deemed irrelevant (i.e., grossly misaligned with the input content). In terms of novelty (given that sometimes simply agreeing with the clinician’s differential was deemed a high-quality response), AI-generated responses (1,384; 94%) provided new diagnostic insights, while a minority (85; 5.8%) offered no novel insight or were misleading. And finally, in terms of contextual relevance (e.g., an awareness of the local epidemiology) in its diagnostic reasoning, nearly all responses (1,384; 94%) were fully aligned with the patient’s clinical and social context, with partial alignment observed in 78 cases (5.3%) and complete misalignment in just 7 cases (0.5%).

#### Clinical Reasoning (Investigations and Treatment Planning)

The vast majority of AI responses (1,460; 99%) provided clinical management advice (e.g., examinations, tests, referrals, treatments). Moreover, almost all AI responses (1,455; 99%) aligned with local clinical guidelines. Furthermore, most AI responses (1,461; 99%) contributed additional management-related insights beyond the initial documentation. And finally, appropriateness of the LLM response in the patient’s socioeconomic or cultural context or resource limitations was high with no conflict in 1,153 encounters (78%); fully adapted in 309 (21%); with only small numbers showing issues - attempted but failed to adapt 2 (0.1%), did not attempt to adapt 2 (0.1%), or not culturally/contextually appropriate 3 (0.2%).

#### Safety Concerns Associated with LLM Outputs

115 responses (7.8%) included active recommendations that evaluators considered potentially harmful: 37 (2.5%) were regarded to have posed major safety concerns and 78 (5.3%) minor concerns. Among these, evaluators judged that in 25 cases (22%), the clinician appeared to have fully adopted the harmful advice, and in 42 cases (37%) partially adopted it. In 48 cases (42%), the harmful recommendations were not acted upon. The types of issues identified in the subset of potentially harmful AI responses were broadly similar to those found in initial clinical documentation.

The most common concerns associated with the LLM outputs were inappropriate medication recommendations (54; 46.2%) and omission of critical differential diagnoses (37; 31.6%). Incorrect diagnoses were noted in 17 responses (14.5%), while less frequent issues included incorrect or unsafe dosing (6; 5.1%), inappropriate investigations (3; 2.6%), culturally/contextually inappropriate guidance (3; 2.6%), and other concerns (24; 20.5%).

### Clinician Response to LLM Recommendations and Impact on Patient Safety

In most encounters (917; 62%) clinicians made no LLM-induced changes; 358 (24%) had minor LLM-induced changes and 194 (13%) had major LLM-induced changes. Among the 552 encounters with any change, edits most often concerned the treatment plan (436; 79%), followed by the differential diagnosis (184; 33%), history or examination findings (131; 24%), and the follow-up plan (129; 23%); investigations were revised in 98 (18%), and other edits occurred in 18 (3%). Of these 552 encounters, the LLM fully mitigated a potential harm in 67 (12.1%) and partially mitigated risk in 51 (9.2%); the remaining 434 (78.6%) either had no initial risk identified or showed no mitigation.

In more than half of all encounters (803; 55%), there was no risk identified initially, and no new safety concerns were introduced. However, in 420 cases (29%), clinicians appeared to continue with the potentially harmful course of action identified in the initial documentation, despite the LLM providing explicitly helpful guidance in 362 cases (25%). Apart from the aforementioned 115 cases of LLM-induced harmful action, there were another 13 cases of spontaneous (not LLM-induced) changes to the final documentation that introduced a risk to the patient (which were not present in the initial documentation) (Figure 3). Figure 4 highlights three (de-identified) real-world examples of LLM-related harm introduction.

**Figure 3:**
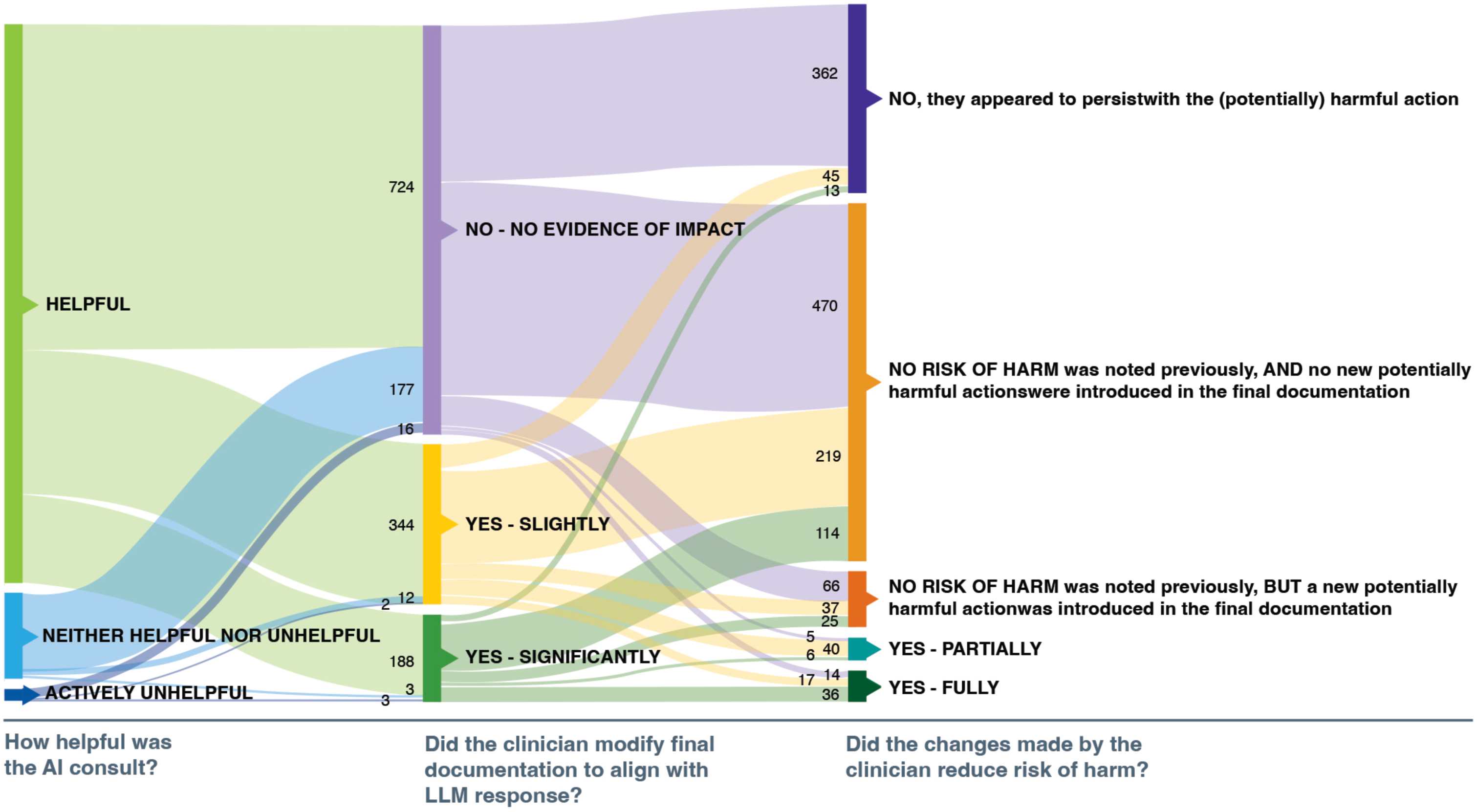
Sankey Plot of Modifications to Final Clinical Documentation, Relationship to LLM Outputs, and Implications for Patient Safety.

**Figure 4:**
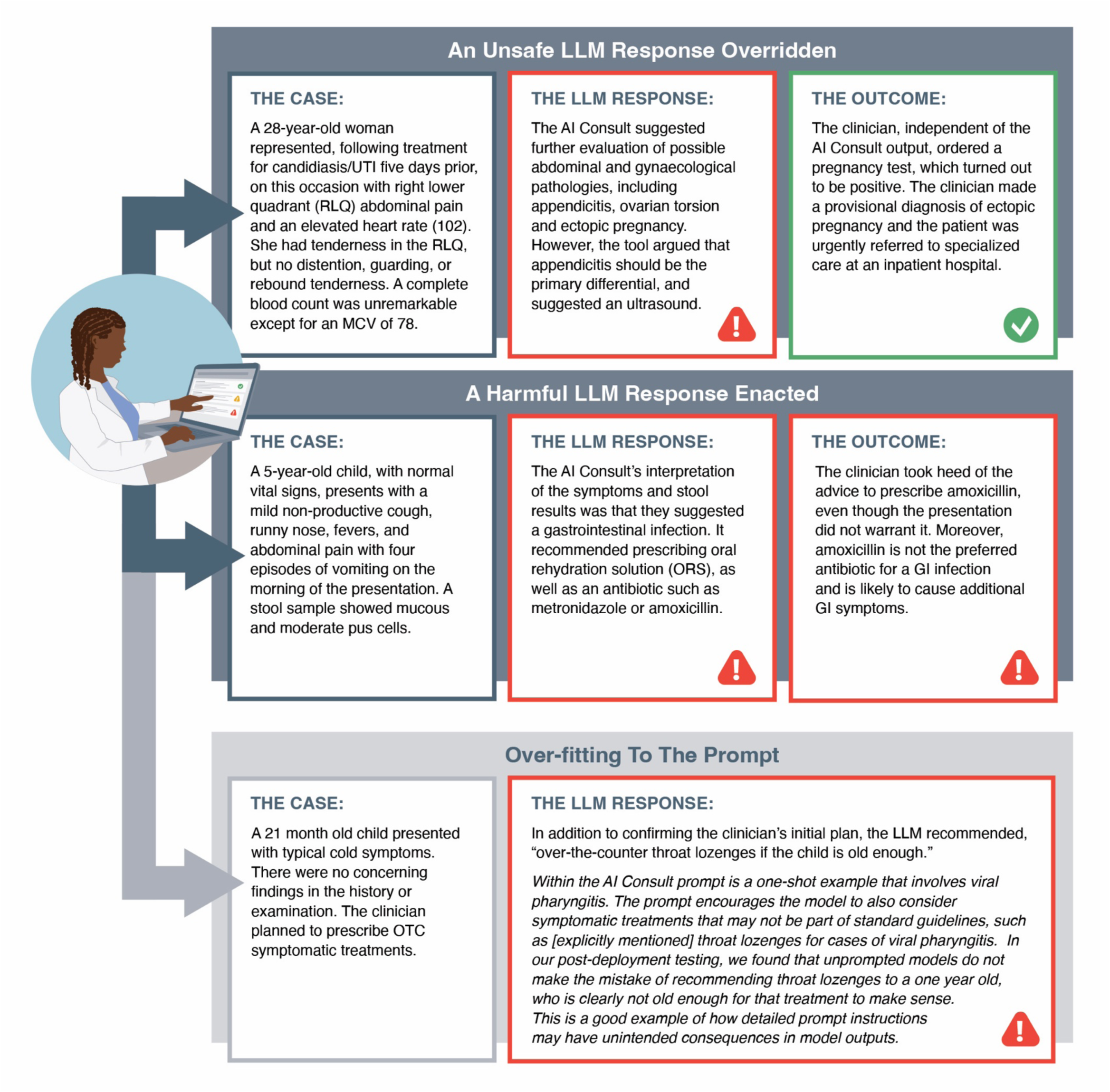
Summaries of Real-World Examples of AI Consult Impacting (or not) Care Quality (Both Positively and Negatively)

## Discussion

In this real-world evaluation of an LLM-based CDSS in Kenyan primary care clinics, we found that the system’s suggestions were largely (clinically) well-reasoned, appropriate to the local context, effectively communicated, and with only a very small number of “hallucinations” (a large number of which were incidental misunderstandings of uncommon acronyms)—all notable given that the underlying system was an off-the-shelf LLM (i.e., not purpose-built, not fine-tuned, and not augmented with contextually relevant information using a Retrieval-Augmented Generation framework). However, in nearly two-thirds of encounters where the LLM was utilized (n = 917, 62%), there was no observable impact on clinician decision-making, suggesting that there is an important difference between interaction/engagement with such tools and incorporation of the outputs. Critically, of these encounters, 362 were instances where the LLM provided beneficial guidance, but the clinician did not heed it, illustrating the unrealized potential of this technology. In instances where the LLM’s advice was acted on (n = 552), approximately twice as many instances of beneficial (to the patient) (n = 118) recommendations were followed compared to harmful recommendations (n = 67). Notably, however, this represents 22% of the instances in which beneficial advice was provided, and 58% of the harmful outputs, suggesting that harmful guidance was much more likely to be acted upon by clinicians. Although the proportion of total encounters (7.8%) that these harmful recommendations reflect is relatively low, it underscores the genuine risks associated with integrating LLMs into care. In summary, the results of the study provide compelling evidence for the potential of LLM-based CDSS to improve the quality of care in low-resource settings, but further research is required to optimize the user experience and utilization of the system outputs, create effective safeguards to reduce the risk of (uncommon) harmful outputs, and confirm the patient-level impact via a controlled, interventional study ^20^.

### Results in the Context of the Literature

Advanced LLMs have shown impressive capabilities on medical exams ^21,22^, and standardized case vignettes ^23^, often matching or surpassing human clinicians on structured clinical problem-solving tasks. This has led to a proliferation of EMR-integrated solutions that leverage these purported clinical reasoning capabilities ^24,25^; however, no other studies appear to report a clinical evaluation of a real-world implementation of an EMR-embedded, CDSS-style copilot, regardless of context, making it difficult to determine whether the results of this study align with expectations. The closest analogue is Google’s OSCE-based (simulated-patient-based) assessment of G-AMIE ^9^, which is an autonomous multi-agent system for information solicitation (the major difference from the AI Consult tool in this study), diagnostic reasoning, and clinical management planning. G-AMIE, based on Google’s Gemini 2.0 Flash, can identify the most appropriate diagnosis and treatment plan in most cases, similar to the abilities observed for AI Consult in this study. Notably, less extensively trained clinicians (nurse practitioners and physician associates), when augmented by the system, appeared to perform better than more extensively trained primary care physicians, largely because the latter group showed higher confidence in their diagnoses and treatment plans despite performing no better, and in some instances worse. In essence, the issue of (unjustified) confidence appears to be a consistent theme in limiting the potential (beneficial) impact of AI-based tools in improving quality of primary care.

A systematic review noted that many AI health care solutions in LMICs remain at the proof-of-concept stage and fully implemented real-world evaluations in low-resource settings are rare, limiting our understanding of their true impact and cost-effectiveness ^14^. Of the limited high-quality evidence available from LMICs (randomized controlled trials), we identified only one economic evaluation of an AI-based tool ^26,27^, and it suggests the technology is not cost-effective ^28^. The total cost of running the LLM-based CDSS for the 1,469 encounters in this study was $7.81, which translates to an average of 0.5 (US) cents, assuming one API call per encounter with an average of 670 input tokens and 364 output tokens. This excludes non-token expenses (engineering/integration, infrastructure, retries, security/compliance, storage, monitoring) but offers a conservative lower-bound on feasibility in low-resource settings. For context, using malaria as a comparator, the willingness to pay in low-resource settings for a rapid diagnostic test is 20 to 50 (US) cents ^29,30^, and for drug therapy is up to $2 ^30^. In essence, the LLM-based CDSS costs are considerably lower than what health systems and individuals (i.e., out-of-pocket expenses) can afford for locally relevant, priority health issues. Whilst we cannot compare efficacy or cost-benefit statistics across the two sets of interventions based on the results of this study, the absolute cost of the proposed AI-based intervention does not appear to be prohibitively large.

As noted earlier, previous reports have cautioned that the deployment of AI models trained predominantly on data from high-income countries may fail to account for local disease epidemiology, resource availability, and cultural practices, thereby limiting their reliability in LMIC environments ^14,31,32^. In this study, we observed some confirmatory evidence of this concern, specifically when the AI’s recommendations were inappropriate, it was often because they were not well tailored to the Kenyan primary care context, such as suggesting a diagnostic test not available in these clinics, or a medication not on local formularies. Assessing solely the absolute number of such events might lead one to believe that this issue is not actually that much of a problem, given that it is substantially smaller than the number of cases in which beneficial recommendations were provided. However, trust in the system overall must be sufficient that clinicians are responsive to the AI advice when it truly matters. Gaining that trust requires consistent demonstration of accuracy and value ^33^. As such, the indirect impact of this minority of cases is that many of the beneficial recommendations were ignored; this was confirmed by a parallel user-experience study conducted with the clinicians at Penda Health, who reported a loss of confidence in the tool when it provided unaligned responses ^18^.

### Strengths & Limitations

A major strength of this study is the multi-site design. By evaluating the CDSS across 16 clinics, we captured how the AI performs in routine urban primary care practice. This breadth improves the relevance and generalizability of our findings within the Kenyan context, compared to simulations or single-site pilots. The study’s evaluation methods engaged an independent panel of experienced local physicians to review cases, with a standardized training and calibration process to improve consistency in their judgments. Multiple reviewers assessed each case domain, and we conducted inter-rater reliability checks to quantify consistency. Additionally, our sample size is substantially larger than most prior evaluations of AI in health care in LMICs, affording more precise estimates of AI performance. Finally, the CDSS was evaluated within the live EMR system, reflecting real clinician–AI interactions with minimal Hawthorne effect, and thus likely capturing authentic behavior under normal time pressure and responsibility.

There are several limitations to this study. First, the observational, retrospective design precludes any definitive conclusions about causality or clinical impact. We did not have a contemporaneous control group of encounters without AI usage, so we cannot quantitatively determine how much the AI actually improved diagnostic accuracy, treatment appropriateness, or efficiency compared to standard care. Any improvements in error rates are based on before-and-after comparisons within the same encounter rather than a randomized comparison across encounters. There is a risk of confounding in cases where clinicians may have selectively used the AI on more complex cases, meaning the overall error profile with AI could differ from that of typical cases for reasons unrelated to the tool itself. A prospective controlled trial is currently underway in Kenya, and its results are expected to provide high-quality evidence on outcomes such as treatment success rates and safety events under LLM-assisted care ^6,20^. Moreover, our evaluation relied on subjective expert review of documentation, introducing some measurement uncertainty. Assessing whether a diagnosis or treatment was appropriate often requires clinical judgement, and experts may differ, especially when documentation is limited. We sought to minimize bias through rater training, explicit criteria, and independent review with adjudication; however, some variability persisted. Notably, of the 37 cases categorized as major safety concerns, 10 were contributed by a single evaluator. In a post-study audit of cases flagged as major, many reflected judgement calls rather than unequivocal errors; most involved omissions (e.g, endorsing an incorrect investigation or treatment already present in the clinician’s initial documentation) rather than the LLM actively recommending a harmful action. (Supplementary Material 3). Finally, the evaluation covered a relatively short time window (three months) and a specific version of the LLM. AI models continue to evolve rapidly, and user behavior also shifts over time. Thus, our snapshot may not capture long-term performance trends, and specifically, the impact of any training effects from repeated nudging.

### Future Research

Several additional avenues of exploration would benefit future implementation programs and product development exercises. Apart from trust, there are likely other factors influencing the suboptimal utilization of LLM outputs, which, if identified, might allow for a better construction of human-computer interaction to increase the uptake of beneficial recommendations and thus improve quality of care. Moreover, the higher likelihood of harmful recommendations being followed than beneficial ones is a related, but equally important, question worthy of further investigation to isolate the cause. Furthermore, future studies might consider exploring different deployment models, such as LLM advice only for certain conditions, only visible to supervising physicians, or limited to serving as training tools and uncoupled from individual clinical interactions, to determine what yields the best balance of benefit and safety.

### Implications for Policy Makers, Product Developers, and Researchers

This study highlights the dependency between technological fixes and systemic infrastructure. The setting of this study was a private, reasonably well-resourced primary care network in an urban African context. The clinicians at Penda Health are supported by a robust cloud-based digital infrastructure, regular training, and have access to relatively comprehensive supplies, laboratories, and imaging capacity. It is this investment in the enabling environment and underlying systems (e.g., supply chains, referral networks, training programs) that meant clinicians could act on the outputs of the AI-based solution. Outcomes and LLM utilization patterns will undoubtedly differ in settings with fewer resources, different patient demographics, or alternative health care workflows ^34^. Caution is therefore warranted in extrapolating our results to dissimilar environments, but this also reflects a broader lesson: technology can only be as effective as the environment allows. Reports from other LMIC deployments have documented practical barriers like poor internet connectivity, lack of device integration, low digital literacy, and poor integration into clinical workflow, limiting the usefulness of AI systems ^35,36^. Our findings reinforce, rather than reduce, the need to address these concerns to ensure equitable benefits.

There are also open questions surrounding the regulatory status of LLM-based technologies in health care settings ^37^. Continuous post-deployment monitoring and evaluation of AI systems should be standard practice, but without a regulatory requirement, it is unclear why developers and deployers would agree to this additional burden. Ongoing surveillance is particularly important given that proprietary AI models are often updated, and performance of any AI tool can drift ^38^. As such, health systems deploying AI should plan for this kind of life-cycle monitoring, similar to pharmacovigilance for new drugs or post-market device surveillance ^39^. The current lack of clarity creates significant inertia and risk associated with progressing products that could be classed as a medical device towards deployment at scale, limiting the ability to realize the benefits of this technology.

## Conclusions

Our findings indicate that EMR-embedded, LLM-based decision support systems have strong potential to enhance clinical decision-making in primary care in sub-Saharan Africa, offering relevant and contextually appropriate guidance in many scenarios. Ongoing clinical trials should confirm whether the observed benefits translate into patient-level impact. Regardless, further research is needed to fully understand how clinicians interact with AI-based CDSS, including when they choose to consult it, how they interpret its advice, and why they sometimes override or ignore it. It appears, at minimum, localizing AI systems (through training on region-specific data and inclusion of national treatment guidelines) will be key to building trust in AI-assisted care, and thus maximizing uptake and therefore impact of this technology. In parallel, interdisciplinary research on governance, ethics, and policy must keep pace with technical advances if we are to fully and equitably unlock the benefits of this technology.

## Methods

### Study Design and Setting

We conducted a retrospective observational study using routinely collected electronic health record data from primary health care facilities in Kenya. The study analyzed data collected at 16 clinics operated by Penda Health, a private social enterprise and primary care network serving urban communities in Nairobi and Kiambu counties. In early 2024, these clinics implemented an LLM-based CDSS integrated into their EMR (described in more detail below and illustrated in Figure 1). The system allows clinicians to click an “AI Consult” button during patient encounters, which prompts the LLM (specifically GPT-4o) to provide actionable diagnostic and therapeutic guidance tailored to the case.

### Participants and Data Collection

We included patient encounters from a three-month inclusion period (July 1, 2024, to September 30, 2024) during which the AI Consult feature was actively used in clinical workflows. Eligible encounters were those in which the attending clinician engaged the AI Consult for the patient’s case using the comprehensive consult prompt. From all such encounters during the period, we drew a proportionally allocated, age-stratified random sample of 1,500 cases (strata: 0–4, 5–17, 18–39, 40– 59, 60+) to preserve the marginal age distribution of the source dataset. This sample was selected for detailed review. Encounters were excluded if the patient had previously opted out of research using their data (in line with the clinic network’s consent policies) and any excluded record was replaced with another randomly selected case from the same period.

All data originated from Penda Health’s EMR system and were extracted from an Azure SQL Server database, which maintains a consolidated replica of the EMR data, separate from the operational system. For each selected encounter, we retrieved de-identified clinical data (patient demographics, presenting complaints, medical history, examination findings, diagnoses, treatments, and follow-up outcomes) alongside the full transcript of the LLM’s consultation (including the input data and the LLM’s output text). A unique study ID was assigned to each record, and any personal identifiers were removed prior to analysis to ensure confidentiality. Data extraction and de-identification were performed by authorized data managers using automated scripts within the secure hospital data environment. No patient contact was required, and all data remained within secure servers during the evaluation.

### LLM CDSS Description

The LLM-based CDSS implemented throughout Penda Health facilities that were evaluated in this study provided decision support by analyzing the patient’s clinical notes and suggesting possible diagnoses, investigations, and management steps. The system operates by the click of a button, where a prompt instructs the LLM to consider the patient’s symptoms, vital signs, and clinical history (entered by the clinician into the EMR) and then generate a consultative response as if advising a primary care physician in an urgent care setting. For example, the LLM output might include a differential diagnosis list, recommendations for additional tests or referrals, and treatment or counseling suggestions. Clinicians could review this output in real-time during the consultation and decide whether to follow or ignore the advice. The LLM (GPT-4o) is a generative model that was pre-trained on a broad medical knowledge base and underwent local testing by the Penda Health clinical team; however, it was not specifically trained on Kenyan patient data prior to deployment. All clinicians received a focused orientation on how to use the tool and its limitations (e.g., that it can produce errors) before its integration into practice. The present study evaluates the historical outputs of this tool in routine use, rather than introducing a new intervention. Notably, the AI Consult tool was first introduced in February 2024, alongside user training.

### Expert Evaluation Panel Selection

Penda Health’s Human Resources department ran an open recruitment following a job advert posted on January 30, 2025 on the organization’s social media pages. A total of 162 applications were received. Eligibility targeted Medical Officers who had completed internship in Kenya, were registered with the Kenya Medical Practitioners and Dentists Council, and were actively practicing at least one hour per week. Screening, including verification of current medical licenses, excluded 116 applicants who did not meet minimum requirements, with 46 candidates advancing. These 46 candidates underwent a curriculum vitae review and a phone interview, after which 30 were selected and received offers. All selected evaluators underwent training on the research protocol and scoring criteria prior to reviewing study cases (see Supplementary Material 2 for the scoring criteria and guidance on completion). This training took the form of a half-day in-person workshop. It included a guided review of example cases (separate from the study sample).

### Calibration and reference standard

After the workshop, a calibration exercise was conducted, during which evaluators scored a set of 30 records (sampled from the 1,500 records). We excluded one routine follow-up visit, resulting in 1,469 for the formal evaluation. Four physicians had independently scored each of these 30 records (authors: AA, RK, SK, and BAM). The panel then held consensus meetings to establish a reference standard for each question. In most items, a single consensus response was reported because either all four reviewers selected it, a simple majority (3 of 4) selected it, or the team discussed and agreed to adopt it. In the rare cases where two responses were shown, the reviewers were evenly split between those two; only those two were considered valid and the ±1 rule did not apply. The ±1 rule provided a narrow tolerance within the same sentiment band. For example, for the options [Yes (strong and well-reasoned); Yes (with some gaps); No (major gaps or misleading); No (irrelevant)] if the consensus was Yes (with some gaps), no No option was admissible under ±1. For three-level items [Yes; Yes (partial); No], if the consensus was No (not at all), no ±1 applied because it was the only valid negative. For four-level scales [Entirely; Mostly; Somewhat; Not at all], if the consensus was Mostly, only the adjacent middle option Somewhat was within ±1; Entirely and Not at all were not. Yes/No questions always had a single valid response with no ±1 tolerance. For risk-of-harm items, consensus was defined with particular care. The ±1 tolerance was applied within severity ranges only (e.g., movement from low to moderate or from severe to life-threatening) and was not applied across the low-to-severe divide. In sum, consensus lay within the sentiment of the agreed response, constrained to adjacent options where specified.

Only those evaluators who demonstrated acceptable agreement with reference standards (defined as no more than 50% discordant but within the ±1 range, as well as no scores two points or more from the reference standard), during this calibration phase were permitted to proceed to rate additional study records, to maintain high inter-rater reliability. Notably, all evaluators passed the calibration exercise.

### Evaluation Procedure

From 1,470 cases in the evaluation dataset, we excluded one routine follow-up visit leaving 1,469 encounters for formal evaluation. There were a total of 1880 LLM-generated responses for the 1469 clinical encounters, indicating that in a minority of cases, the AI Consult tool was used more than once in a clinical encounter. In this study, we only consider the initial AI Consult response in relation to the case. Following calibration, the evaluation panel independently reviewed each LLM-assisted encounter. Cases were grouped into batches of 30, with a maximum of six batches per evaluator to limit fatigue and potential scoring bias; each evaluator reviewed 2 to 4 batches.

Each record in the study sample was reviewed by at least one evaluator. To monitor quality and reliability, a random 10% subset of the cases was independently evaluated by a second panelist, who was blinded to the first reviewer’s scores. This enabled the calculation of inter-rater reliability metrics and helped identify any systematic inconsistencies. The evaluators performed their reviews using a secure online portal where the anonymized records were assigned. They were instructed to rely on standard Kenyan clinical practice guidelines (which were made available via a reference repository) and their clinical judgment when assessing the appropriateness of the AI’s suggestions. They were not permitted to discuss cases with each other to avoid influence, but they could query the study coordinators or investigators if any patient record data was unclear.

For any case where the AI’s advice included potentially harmful elements (such as a dangerous medication error or an unsafe management plan), the evaluator documented this finding. In such instances, an additional review was performed: the patient’s follow-up records and any safety incident reports were examined (by the Penda Health clinical quality team) to determine if the clinician had actually acted upon the questionable AI recommendation and whether it led to any adverse outcome. Any such events were then managed in accordance with Penda Health’s standard operating procedures for clinical safety events, and reporting was deemed to be beyond the scope of this study.

### Outcomes and Measures

The primary outcomes of interest were:

1. The quality of the initial documentation;
2. The appropriateness of any diagnostic reasoning;
3. The appropriateness of any clinical reasoning (including proposed investigations and treatment planning); and,
4. The safety of the LLM’s recommendations, and whether both potentially harmful and beneficial recommendations appeared to be acted upon.

Results from the questions aligned with the above outcomes were summarized. For cases with identified unsafe advice, we recorded the nature of the risk (e.g., misdiagnosis, incorrect medication, etc.) and whether any actual patient harm occurred (as evidenced by follow-up data).

### Statistical Analysis

We used descriptive statistics to summarize the data. Variables were reported as counts and percentages. Inter-rater reliability was assessed on the subset of duplicatively assessed cases, each independently reviewed by two evaluators. For ordinal outcomes, including the quality of initial clinical documentation, whether the LLM addressed identified issues, alignment with patient context, clinician modification of documentation, and reduction of identified risks, we calculated Kendall’s W statistic to quantify agreement among reviewers. For binary and nominal outcomes, including the presence of unsafe advice, hallucinations, evidence of diagnostic reasoning, and provision of a reasonable differential diagnosis, we calculated Fleiss’ kappa. Agreement strength was interpreted using the Landis and Koch benchmarks ^40^. All analyses were conducted using R (version 4.5.1).

### Ethical Considerations

This study was conducted in accordance with international and national ethical guidelines for research. The protocol was reviewed and approved by the Amref Ethics and Scientific Review Committee (ESRC) in Kenya (P1839-2025). Given that the research involved secondary use of de-identified clinical data and posed minimal risk to patients, a waiver of informed consent was obtained from the ethics committee. To protect patient confidentiality, all data were anonymized before analysis, and no personal identifiers appear in any reports or outputs. The evaluation panel members signed confidentiality agreements and accessed the records through a secure platform. All procedures adhered to the principles of the Declaration of Helsinki and Good Clinical Practice.

## Data Availability

De-identified quantitative data are available from the corresponding author, for replication of the study results, upon reasonable request, and will be subject to the original Amref ESRC (the institutional ethics review board) approvals/restrictions. All further analyses will require agreement with the clinical partner (Penda Health), and separate approvals from a local ethics review board in line with best practices for the unconsented reuse of routine data.

## Declarations

### Patient and Public Involvement Statement

Patients were not directly involved as research participants.

### Author contributions (CRediT taxonomy)

Conceptualization: BAM; Methodology: BAM, MEF, RK, AA, PM, WM, SK; Software (EMR integration and AI Consult implementation): RK, NA; Data curation (extraction, de-identification, management): NA, RK, PM, WM; Investigation: all authors; Formal analysis: PM; Visualization: PM, BAM; Project administration: MEF, WM, SK, LI, CW, NA; Supervision: BAM, AA, RK; Funding acquisition: BAM, MEF; Writing (original draft): AA; Writing (review and editing): all authors

## Acknowledgments

We thank the experts who served on the evaluation panel: Zeitun Mohamed Abey; Kerry Atebe; Agnes Chepkorir; Elizabeth Khanali Chesoni; Antony Onyango Clinton; Zinzi Gecheo; Paul Gitonga; Eliash Dawn Jakech; Jackline Kaharo; Ann Nduta Kamanda; Lucky Kiguongo; Wachira Wilson Kimunya; Maureen Wanjiru Kinuthia; Wallace Kipleting; Ann Monicah Wambui Kiragu; Melvin Wanjala Masibayi; Ian Muchiri; Jason Muga Muchira; Sylvia Vusevwa Mugomangi; Effie Faith Mwalo; Peter Maiya Ndubi; Angela Ngugi; Beverly Bioreri Nyarigoti; Okangi George Obeya; Patricia Sapiri Omedo; Annette Marcella Onyango; Austine Ochieng Ouma; Antony Owaya; Kevin Tony Okoth Oyugi; and Rachel Melissa Salins.

## Supplementary material

Supplementary Material 1: sBox, sTables and sFigures

sBox 1: Full wording of the comprehensive AI consult prompt used in the evaluation.

sTable 1: Inter-rater reliability statistics across evaluated metrics.

sTable 2: Likelihood, severity, and type of harm by initial documentation quality – all records (n = 1,469), of which 547 (37%) prompted safety concerns.

sTable 3: Likelihood, severity, and type of harm by initial documentation quality – high-quality documentation (n = 250), of which 36 (14%) prompted safety concerns.

sTable 4: Likelihood, severity, and type of harm by initial documentation quality – acceptable documentation (n = 878), of which 309 (35%) prompted safety concerns.

sTable 5: Likelihood, severity, and type of harm by initial documentation quality – needs significant improvement (n = 279), of which 163 (58%) prompted safety concerns.

sTable 6: Likelihood, severity, and type of harm by initial documentation quality – totally inadequate documentation (n = 62), of which 39 (63%) prompted safety concerns.

Supplementary Material 2: Evaluator Question and Response Rubric

Supplementary Material 3: Event Summaries and Study Team Review of Each ‘Major Safety Concern’ Event Associated with LLM Output

